# Association between Prenatal Adversity and Neonatal Outcomes in Infants with Prenatal Opioid Exposure

**DOI:** 10.64898/2026.10.08.26365058

**Authors:** Madison Ramirez Heil, Macie Donahue, Marie Camerota, Lynne Dansereau, Alexandrea Craft, Elisabeth Conradt, Barry Lester

## Abstract

**Objective:** Examine whether prenatal adversity is associated with Neonatal Opioid Withdrawal Syndrome (NOWS) severity and the need for pharmacological treatment in infants with prenatal opioid exposure.

**Study Design:** Prospective cohort study of 235 infants from a multi-center study of infants with prenatal opioid exposure. Maternal factors were assessed using the electronic medical record and maternal questionnaires. Latent Class Analysis (LCA) was used to derive mutually exclusive prenatal adversity groups.

**Results:** A 3-class LCA solution provided the best fit to the data and showed 3 distinct groups: one low adversity group (Class 3, 38.7%) and 2 high adversity groups. Infants born to mothers with elevated demographic, medical, polysubstance, and mental health risk factors (Class 1, 31.9%) were more likely to require pharmacological treatment for NOWS (OR=7.70, 95% CI: 3.65-17.14) and had higher NOWS severity, as indexed by longer length of stay (β = 6.08, 95% CI: 4.09-8.07), and higher maximum Finnegan scores (β = 2.62, 95% CI: 1.49-3.76). Infants born to mothers with elevated medical and polysubstance risk factors (Class 2, 29.4%) were also more likely to receive pharmacological treatment for NOWS (OR=3.22, 95% CI: 1.56-6.78) and had higher NOWS severity, as indexed by higher likelihood of receiving adjunctive medication (OR=3.62, 95% CI: 1.11-12.99).

**Conclusion:** Maternal sociodemographic, mental health, substance use, and medical adversity may serve as an early marker of NOWS severity and may aid in the prediction of which neonates will require pharmacological treatment for NOWS. Early identification of infants at high risk for NOWS could lead to earlier treatment and the potential to reduce NOWS severity, length of stay, and associated healthcare costs.

## Introduction

The opioid epidemic has profoundly impacted maternal and infant health, with increasing opioid use during pregnancy accompanied by a rise in Neonatal Opioid Withdrawal Syndrome (NOWS). From 2010 to 2017, maternal opioid-related diagnoses increased from 3.5 to 8.2 per 1,000 delivery hospitalizations while the rate of NOWS increased from 4.0 to 7.3 per 1,000 birth hospitalizations [1].

Maternal opioid-related diagnoses are also associated with co-occurring mental and physical health conditions [2,3]. Thus, prenatal opioid exposure often occurs within a broader context of maternal health and adversity that may shape risk to both mothers and their infants.

Within this context, the clinical presentation of NOWS varies considerably depending on type and degree of opioid exposure, but also other factors such as maternal and infant metabolism and co-occurring, non-opioid exposures (e.g., cigarettes, anxiolytics) [4]. Although maternal and prenatal factors are known to impact the clinical presentation of NOWS, less is known about how multiple prenatal adversities may impact NOWS severity and outcomes [4–6]. This gap is notable as women who use opioids during pregnancy may be more likely to experience sociodemographic, psychological, and medical risk factors that impact both maternal and infant health [7,8]. Several of these individual risk factors such as maternal depression, antidepressant use and maternal weight have been associated with greater NOWS severity, including longer length of treatment and more severe withdrawal symptoms as reflected by higher Finnegan scores [8–10]. However, examining these factors separately may not capture clinically meaningful patterns of co-occurring maternal risk.

Given the prevalence and co-occurrence of prenatal adversity among women who use opioids during pregnancy, understanding patterns of maternal risk may help characterize infants at greater risk for NOWS. Yet few predictive models exist to determine NOWS onset or severity [11]. The objective of this study is to examine whether patterns of maternal prenatal adversity are associated with NOWS severity and the need for pharmacological treatment among infants with prenatal opioid exposure.

## Methods

### Participants

Participants are from the Child and Family Study (CAFS), a multi-site, prospective study seeking to distinguish novel clinical predictors of NOWS. Between 2019 and 2026, 235 mother-infant dyads with prenatal opioid exposure were recruited postnatally and from prenatal clinics at Women and Infant’s Hospital of Rhode Island and University of Utah Hospital. Mothers were approached for consent if they attended an outpatient specialty clinic for women with opioid use and other substance use disorders (Utah site) or if prenatal opioid use was identified via maternal medical record during pregnancy or at delivery, a positive infant umbilical cord or urine toxicology screen after birth, and/or a positive maternal urine toxicology screen during pregnancy or at hospital admission (Rhode Island site). Exclusion criteria included gestational age <33 weeks, inability to take oral medications, metabolic disturbances, congenital anomalies, genetic syndromes, asphyxia, seizures, respiratory failure, or sepsis. Caregivers who were unable to provide informed consent or infants not medically stable for a newborn neurobehavioral examination were also excluded. Study procedures were approved by the Institutional Review Board of each study site. All caregivers provided written informed consent for participation.

### Prenatal Adversity

Maternal sociodemographic, mental health, substance use, and medical risk factors were collected via a combination of medical record review and maternal questionnaires. Sociodemographic factors included maternal age >35, income <100% poverty line, education less than high school degree, minoritized race or ethnicity (i.e., race or ethnicity other than non-Hispanic white), and lack of a relationship partner. Mental health risk factors included anxiety and/or depression during pregnancy, antidepressant, antianxiety, or antipsychotic medication use, Post Traumatic Stress Disorder (PTSD), bipolar disorder, and elevated emotion dysregulation, defined as scores above the 75^th^ percentile on the Difficulties in Emotion Regulation Scale (DERS)[12] in our sample. Maternal substance use variables included alcohol, tobacco, marijuana, and non-opioid illicit drug use during pregnancy. Medical risk factors included inadequate prenatal care, pre-pregnancy underweight (BMI < 18.5), pre-pregnancy obesity (BMI ≥ 30), pregnancy weight gain > Institute of Medicine guidelines [13] based on pre-pregnancy BMI and reported pregnancy weight gain, hypertension, pre-eclampsia, diabetes, and any sexually transmitted disease (STD).

### NOWS Treatment and Severity and Treatment

Measures of NOWS severity and pharmacological treatment were abstracted from the infant’s medical record. Criteria for initiation of pharmacological treatment for NOWS was based on study site hospital practices. The Finnegan Neonatal Abstinence Scoring Tool (FNAST) was used at Women and Infants Hospital [14] . The Neonatal Withdrawal Inventory (NWI) and Eat Sleep Console (ESC) were used at the University of Utah Hospital [15,16]. To account for variability in pharmacological treatment rates due to different assessments, study site was adjusted for in the analyses. Both sites used morphine as the first line of pharmacological treatment to treat infants meeting criteria for NOWS. For infants requiring adjunctive medication, phenobarbital was the second line of pharmacological treatment. NOWS severity was indexed by length of treatment (LOT), length of hospital stay (LOS), maximum FNAST score, maximum morphine dose, and need for adjunctive medication.

### Statistical analysis

To examine patterns of prenatal adversity, we applied latent class analysis (LCA) to our set of 25 prenatal risk factors. LCA uses maximum likelihood estimation to categorize individuals into mutually exclusive subgroups based on patterns of responses on a number of categorical variables. This method enabled us to study subgroups of mothers with similar, co-occurring prenatal risk factors. We fit LCA models with 1 to 4 classes and examined model fit statistics to identify the best fitting model. All models were run in Mplus 8.11 and full information maximum likelihood was used to handle missing data. We used posterior probabilities to group participants into their most likely adversity subgroups. For descriptive purposes, we used binary logistic regression with pairwise comparisons to test differences between the subgroups on the 25 prenatal adversity variables. To test associations between prenatal adversity and NOWS outcomes, we used linear and logistic regression models, for continuous and categorical outcomes, respectively. Covariates included study site and infant sex. Additional demographic factors were not included as covariates because they were included as part of the LCA analysis.

### Results

The CAFS study included 235 infants born to 227 mothers (**Table 1**). The mean GA of infants was 38.3 weeks (SD=1.7 weeks), 51% of the infants were male, and Department of Children, Youth, and Families (DCYF) was involved for 49% of infants. Of the 227 mothers, 45% did not have a partner, 25% did not complete high school, and 21% were in residential treatment for opioid use. In addition, 21% had hypertension and 31% had Hepatitis C.

**Table 1.** Maternal and Infant Characteristics.

|  | M (SD) or N (%) |
| --- | --- |
| **Maternal Characteristics (N=227)** |  |
| Less than high school degree | 56 (24.7%) |
| Inadequate prenatal care | 40 (17.6%) |
| No partner | 102 (44.9%) |
| Hypertension | 47 (20.7%) |
| Residential treatment | 47 (20.7%) |
| Inpatient detox during pregnancy | 41 (18.1%) |
| Bipolar | 50 (22.0%) |
| PTSD | 83 (36.6%) |
| Hepatitis C | 70 (30.8%) |
| **Infant Characteristics (N=235)** |  |
| Sex (Male) | 120 (51.1%) |
| Gestational Age (weeks) | 38.3 (1.7) |
| Birth Weight (grams) | 3016 (543) |
| Head circumference at birth (cm) | 33.8 (2.2) |
| Length at birth (cm) | 48.4 (3.2) |
| DCYF Involvement | 114 (48.5%) |
| Breastfeeding | 138 (58.7%) |

### Prenatal adversity subgroups

We fit LCA models with 1 to 4 classes. Model fit statistics are shown in **Table 2**. A 3-class LCA solution provided the best fit to the data and had the lowest Bayesian Information Criterion (BIC), high entropy (0.85) and high class probabilities (0.90-0.96) and showed 3 distinct subgroups: one low adversity group (Class 3, 38.7%) and 2 high adversity groups (Class 1, 31.9% and Class 2, 29.4%; **Figure 1**). Women in the three latent classes differed on 18 of the 25 prenatal risk factors (**Table 3**).

**Table 2.** Model fit statistics for LPA models

| Number of profiles | Convergence problems | Lowest loglikelihood replicated | BIC | Entropy | Average Class Probabilities | Smallest Class Size | LMRT *p*-value | BLRT *p*-value |
| --- | --- | --- | --- | --- | --- | --- | --- | --- |
| 1 | No | Yes | 6232 | -- | -- | -- | -- | -- |
| 2 | No | Yes | 6089 | .748 | .90 - .93 | N =102 (43%) | 0.075 | < .001 |
| 3 | No | Yes | 6084 | .845 | .90 - .96 | N = 69 (29%) | 0.169 | < .001 |
| 4 | Yes | No | -- | -- | -- | -- | -- | -- |

*Note.* BIC = Sample size adjusted Bayesian Information Criterion; LMRT = Lo-Mendell-Rubin test; BLRT = Bootstrapped likelihood ratio test.

**Table 3.** Distribution of prenatal risk factors by latent class

| **Prenatal Risk Factors** | **Class 1<sup>a</sup>** | **Class 2<sup>b</sup>** | **Class 3<sup>c</sup>** | ***p*-value** | **C1 vs C2** | **C1 vs C3** | **C2 vs C3** |
| --- | --- | --- | --- | --- | --- | --- | --- |
| Maternal Age > 35 Years | 19.20% | 41.80% | 37.20% | 0.009 | 0.003 | 0.010 | 0.565 |
| < 100% Poverty Line | 70.50% | 54.10% | 40.00% | 0.010 | 0.125 | <0.001 | 0.182 |
| Education < High School | 24.70% | 33.30% | 19.30% | 0.139 | 0.259 | 0.416 | 0.051 |
| Race Other Than Non-Hispanic White | 7.50% | 4.90% | 7.70% | 0.784 | 0.548 | 0.958 | 0.498 |
| No Partner | 63.50% | 47.10% | 31.10% | <0.001 | 0.046 | <0.001 | 0.040 |
| Inadequate Prenatal Care | 20.00% | 33.30% | 5.60% | <0.001 | 0.072 | 0.006 | <0.001 |
| Pre-pregnancy Underweight | 4.20% | 14.50% | 4.70% | 0.037 | 0.042 | 0.885 | 0.051 |
| Too Much Weight Gained | 57.70% | 47.50% | 34.90% | 0.018 | 0.239 | 0.004 | 0.127 |
| Pre-pregnancy Obesity | 19.70% | 14.50% | 29.40% | 0.084 | 0.424 | 0.156 | 0.025 |
| Hypertension | 24.00% | 25.40% | 15.40% | 0.234 | 0.850 | 0.166 | 0.126 |
| Pre-eclampsia | 9.30% | 6.00% | 8.80% | 0.735 | 0.448 | 0.904 | 0.496 |
| Diabetes | 8.00% | 7.50% | 9.90% | 0.845 | 0.905 | 0.669 | 0.588 |
| Any STD | 56.10% | 32.30% | 26.60% | <0.001 | 0.005 | <0.001 | 0.464 |
| Alcohol | 13.30% | 15.90% | 6.60% | 0.154 | 0.658 | 0.152 | 0.068 |
| Illicit Drugs (non-opioids) | 85.30% | 100.00% | 4.40% | <0.001 | <0.001 | <0.001 | <0.001 |
| Nicotine | 78.70% | 81.20% | 52.70% | <0.001 | 0.709 | <0.001 | <0.001 |
| Marijuana | 62.70% | 69.60% | 1.10% | <0.001 | 0.380 | <0.001 | <0.001 |
| Maternal Depression During Pregnancy | 47.90% | 4.50% | 22.50% | <0.001 | <0.001 | <0.001 | <0.001 |
| Maternal Anxiety During pregnancy | 53.40% | 7.50% | 12.40% | <0.001 | <0.001 | <0.001 | 0.302 |
| Bipolar | 50.70% | 9.10% | 8.90% | <0.001 | <0.001 | <0.001 | 0.965 |
| PTSD | 70.70% | 36.40% | 8.90% | <0.001 | <0.001 | <0.001 | <0.001 |
| Any Antidepressant | 84.00% | 8.70% | 34.10% | <0.001 | <0.001 | <0.001 | <0.001 |
| Any Antianxiety | 82.70% | 33.30% | 31.90% | <0.001 | <0.001 | <0.001 | 0.845 |
| Any Antipsychotic | 52.00% | 8.70% | 5.50% | <0.001 | <0.001 | <0.001 | 0.440 |
| High Difficulties in Emotion Regulation Scale (>75 percentile) | 47.90% | 17.20% | 16.50% | <0.001 | <0.001 | <0.001 | 0.908 |

<sup>a</sup> Demographic, polysubstance, and mental health risk, <sup>b</sup> Polysubstance and increased medical risk, <sup>c</sup> Low Risk

**Figure 1.**
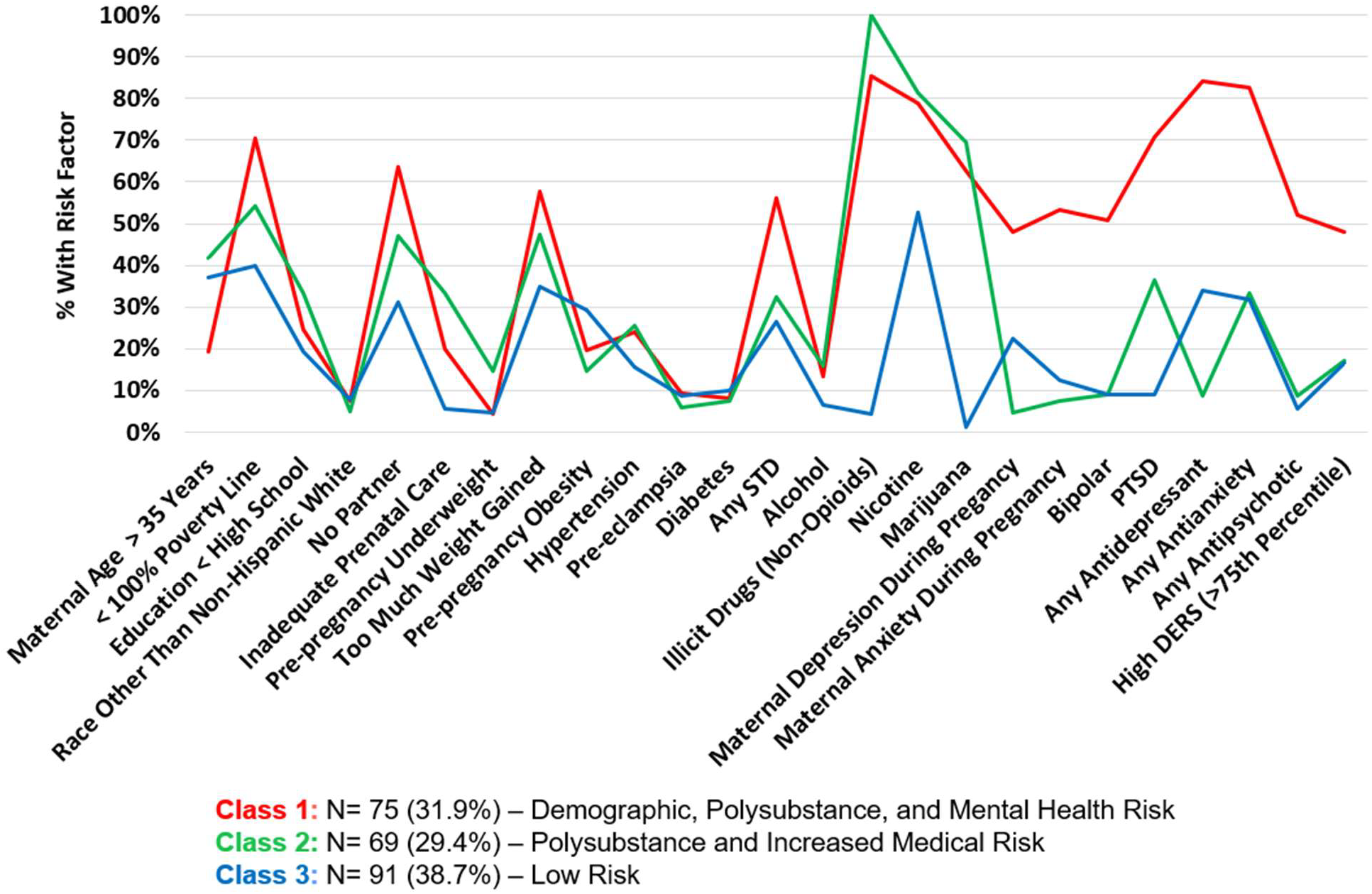
Latent Class Analysis of Prenatal Risk Factors.

One of the high adversity groups (Class 1) was comprised of women with elevated demographic, medical, polysubstance, and mental health risk factors. They were younger moms, with increased poverty, who were unlikely to have a relationship partner. They gained more weight during pregnancy and were more likely to have a sexually transmitted disease. They had higher rates of prenatal alcohol, nicotine, marijuana and illicit drug use and had the highest rates of maternal psychopathology including high rates of bipolar disorder (51%) and PTSD (71%).

The second high adversity group (Class 2) was comprised of women with elevated medical and polysubstance risk factors. They had the highest rates of underweight pre-pregnancy and inadequate prenatal care. They had similarly high rates of prenatal alcohol, nicotine and marijuana use as in Class 1, and the highest rates of illicit drug use of all three groups.

The third class (38.7%) was comprised of women with the lowest overall prenatal adversity. They were more likely to have a relationship partner and adequate prenatal care. They also had the lowest rates of prenatal alcohol, nicotine, marijuana, and illicit drug use and the lowest or similarly low rates of maternal psychopathology as compared to Class 2.

### Prenatal adversity and NOWS

Compared to infants born to mothers in Class 3, infants born to mothers in Class 1 were more likely to require pharmacological treatment for NOWS (OR=7.70, 95% CI: 3.65-17.14; **Table 4**) and had higher NOWS severity, as indexed by longer LOS (β = 6.08, 95% CI: 4.09-8.07), and higher maximum Finnegan scores (β = 2.62, 95% CI: 1.49-3.76). Infants born to mothers in Class 2 were also more likely to receive pharmacological treatment for NOWS (OR=3.22, 95% CI: 1.56-6.78) and had higher NOWS severity, as indexed by higher likelihood of receiving adjunctive medication (OR=3.62, 95% CI: 1.11-12.99) compared to infants born to mothers in Class 3. As compared to Class 2, infants born to mother in Class 1 were also more likely to require pharmacological treatment for NOWS (OR = 2.39 95% CI: 1.15-5.12).

**Table 4.**
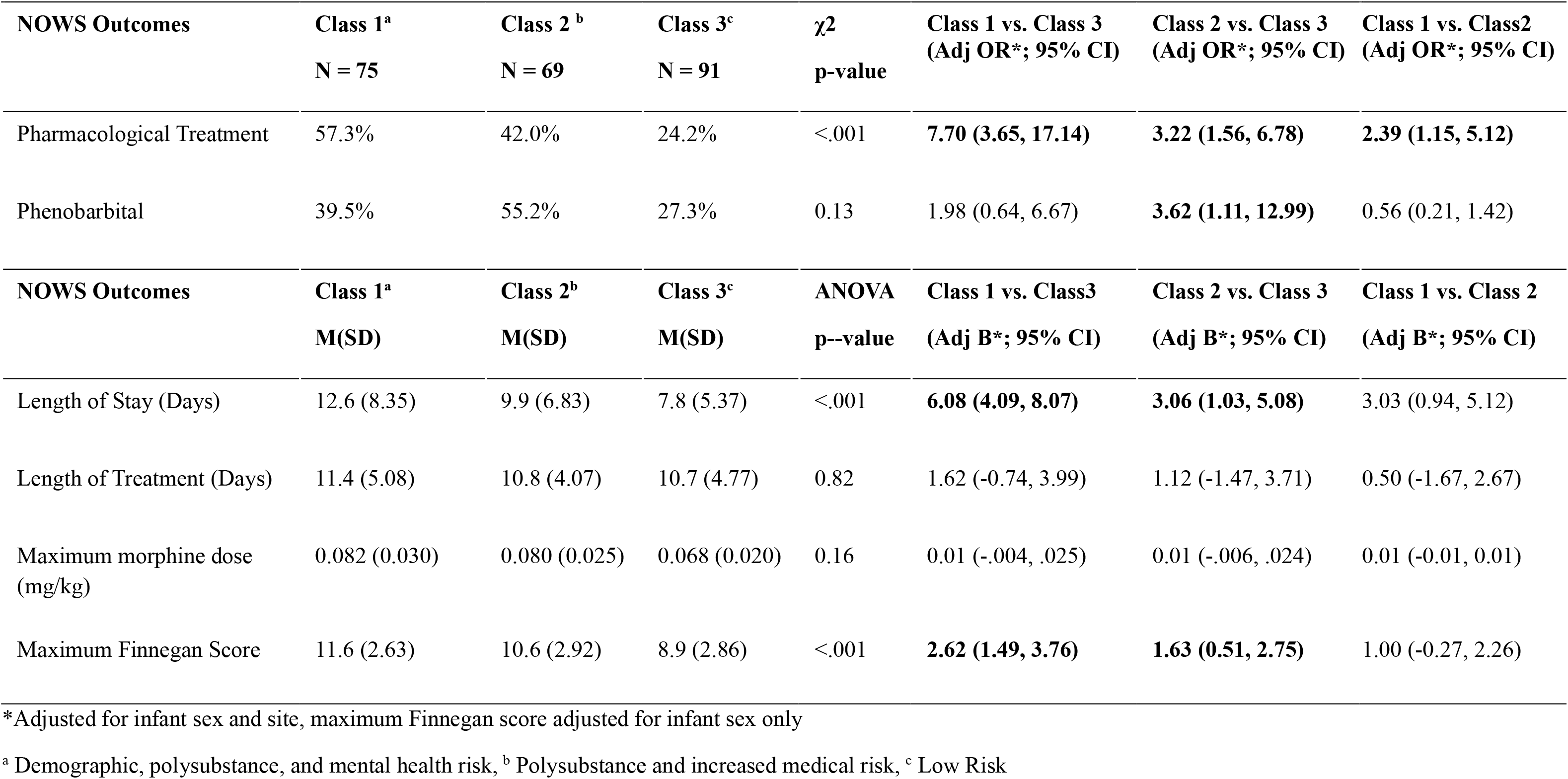
NOWS Outcomes by Prenatal Risk Classes.

## Discussion

The objective of this study was to investigate whether patterns of prenatal adversity as assessed by maternal sociodemographic, substance use, mental health, and medical risk factors was associated with NOWS severity and the need for pharmacological treatment among infants with prenatal opioid exposure. We found that mothers with elevated demographic, medical, polysubstance, and mental health risk factors had infants who were more likely to require pharmacological treatment and higher NOWS severity.

Our latent class analysis identified three distinct patterns of maternal prenatal risk, highlighting the heterogeneity among pregnancies affected by maternal opioid use. Class 1 and Class 2 were comprised of approximately 60% of the sample and represented the higher prenatal adversity groups, distinguished by the specific types of risk factors experienced. Class 3 was comprised of approximately 40% of the sample and represented a comparatively lower adversity profile. These findings suggest that maternal risk among pregnancies affected by opioid exposure is heterogeneous, multidimensional, and may be better characterized by patterns of co-occurring risk factors rather than by individual exposures considered in isolation. In fact, previous studies have shown that women with psychiatric disorders are more than twice as likely to have Substance Use Disorders (SUD) when compared to women without psychiatric disorders [17]. Psychiatric comorbidity, depression and anxiety being the most common, are estimated to affect approximately 25-33% of this population [2]. Our study confirms these findings as mothers in Class 1 were characterized by higher mental health risk with higher probabilities of depression, anxiety, bipolar disorder, PTSD, psychiatric medication use, and difficulties in emotion regulation. This finding is clinically significant because it shows that substance use does not happen in isolation and other risk factors, such as mental health, should be factored into the clinical care for mothers using opioids and their infants.

In contrast, Class 2 was characterized by polysubstance exposure in conjunction with greater medical risk. These findings are consistent with previous studies that showed women who use opioids during pregnancy are more likely to have a lower body mass index (BMI) [7]. Additionally, between 2016 and 2019, women who reported prenatal use of any opioid were less likely to receive adequate prenatal care compared with those who did not report use of opioids [18]. The distinction between Class 1 and Class 2 is notable because mothers with substantial polysubstance use did not constitute a homogeneous group; rather, medical and psychiatric risk clustered differently across latent classes. These results thus emphasize the importance of using a person-centered approach to identify clinically meaningful subgroups that may be missed when studying prenatal risk factors individually.

These adversity classes were also clinically meaningful in their relation to NOWS outcomes. Most notably, infants born to mothers in Class 1 and Class 2 were significantly more likely to require pharmacological treatment for NOWS and have higher NOWS severity, indexed by higher maximum Finnegan scores and longer length of stay as compared to Class 3. Furthermore, even among the two higher adversity groups, infants born to mothers in Class 1 appeared to have the highest NOWS severity and need for pharmacological treatment. Class 1 thus describes a subgroup in which multiple overlapping psychosocial, mental health, and substance use vulnerabilities, experienced together, are associated with greater clinical expression of neonatal withdrawal, beyond what is captured in Class 2 with polysubstance exposure and medical risks alone. While many studies have linked certain individual or potentially predictive risk factors to NOWS outcomes [6,8–10], our findings suggest that the severity of neonatal withdrawal may be influenced by the cumulative and co-occurring nature of maternal risk factors rather than differences in opioid exposure alone. Our findings further validate the need for holistic approaches to care in this vulnerable population.

These findings should be interpreted considering the following limitations. Because this is an observational study, we can identify links between prenatal adversities and NOWS outcomes, but these findings do not necessarily indicate that prenatal adversities are causally related to more severe NOWS. Additionally, although the use of a latent class approach allows for a holistic, person-centered approach to understanding opioid use in the context of other types of prenatal adversity, this approach does not shed light on which specific prenatal risk factors increase the risk for poor NOWS outcomes. While prior research related to prevalence of the prenatal risk factors we assessed align with our findings, our study was comprised of only two hospital sites and therefore may not be fully representative of the greater population of mothers who use opioids during pregnancy. Larger studies with greater geographic representation are needed to investigate the generalizability of these findings.

Due to the variability in NOWS, improved clinical tools to predict risk and guide treatment for infants with prenatal opioid exposure could allow for more individualized and efficient treatment. The current study suggests that considering prenatal risk factors holistically in addition to opioid exposure may enhance prenatal care and aid in the identification of infants at high risk for more severe NOWS. Additional studies may add to our findings and improve the clinical utility of a predictive model of care for infants with prenatal opioid exposure.

## Data Availability

Due to the sensitive nature of our participant's data, all data produced in the present study are available upon reasonable request to the authors.

## Abbreviations

(NOWS): Neonatal Opioid Withdrawal Syndrome
(LCA): Latent Class Analysis
(PTSD): Post Traumatic Stress Disorder
(BIC): Bayesian Information Criterion

